# Longitudinal and Cross-Sectional Associations Between Emotion Recognition Bias, Cognitive Abilities, and Mental Health in the ALSPAC Cohort

**DOI:** 10.1101/2025.10.28.25338883

**Authors:** Rumeysa Kuruoğlu, Robyn E. Wootton, Paul Madley-Dowd, Ian S. Penton-Voak

## Abstract

**Background:** Depression and anxiety are associated with negative emotion recognition biases and cognitive impairments. Better understanding of these relationships could inform potential interventions.

**Aims:** To examine the cross-sectional and bidirectional longitudinal relationships between depression, anxiety and wellbeing with emotion recognition bias, working memory and visual memory.

**Methods:** We examined these relationships in the Avon Longitudinal Study of Parents and Children (ALSPAC) using data from ages 24 and 31-32 years (N = 3991) for longitudinal analysis and ages 31-32 for cross-sectional analysis. We ran linear regressions and reported models unadjusted and adjusted for baseline confounders.

**Results:** Cross-sectionally, higher depressive and anxiety symptoms were associated with negative emotion recognition bias (*b*= −.025, 95%CI[−.042, −.009], *p*= .003; *b*= −.019, 95%CI[−.036, −.003], *p*= .023) but not with working memory errors (*b*= .006, 95%CI[−.023, .034], *p*= .700; *b*= −.001, 95%CI[−.030, .028], *p*= .942), and visual memory (*b*= .009, 95%CI[−.039, .014], *p*= .658; *b*= .008, 95%CI[−.029, .045], *p*= .658).

Longitudinally, depression predicted negative emotion recognition bias in later years (*b*= −.362, 95%CI[−.630, −.094], *p*= .008). There was no evidence that depression predicted higher working memory errors or lower visual memory performance (*b*= .105, 95%CI[−.344, .554], *p*= .645; *b*= −.024, 95%CI[−.598, .549], *p*= .933). Lower accuracy in recognising happy faces predicted future depression (*b*= −.176, 95%CI[−.274, −.079], *p*< .001), but not sad faces (*b*= .073, 95%CI[−.027, .174], *p*= .152) and overall accuracy (*b*= −.012, 95%CI[−.041, .016], *p*= .388). No differences were found between remitted and current depression in emotion recognition bias or cognitive abilities (*p*> .05).

**Conclusions:** These findings suggest that negative emotion recognition bias may be both a symptom and a risk factor for depression, with cross-sectional and longitudinal associations. Cross-sectionally, it appears more linked to general mental health than depression alone. While cognitive abilities show no strong relationship with mental health.

## Introduction

Depression is a debilitating mental health condition that affects millions of people worldwide (World Health Organization, 2017). Identifying key areas for targeted interventions is essential to address this widespread issue effectively. Young adulthood is a critical period for such interventions, as the early onset of depression has been shown to predict depression in later adulthood and depression frequently emerges during this life-stage (McGrath et al., 2023; Park et al., 2014; Solmi et al., 2022). Research suggests that depression is associated with negative biases in facial emotion recognition and impairments in cognitive abilities (Dalili et al., 2015; Rock et al., 2014). Investigating these relationships further is important, as they may serve as potential intervention targets for depression (Pringle et al., 2011; Warren et al., 2015). Depression and anxiety are often comorbid (Essau et al., 2018), and well-being is a key indicator of general mental health and depressive symptoms (Edmondson & Macleod, 2014). Investigating anxiety and well-being alongside depression is important to help clarify whether cognitive impairments and biases are specific to depression or more broadly associated with poor mental health.

Emotion recognition bias is a well-documented social-cognitive bias, where depressed individuals tend to perceive emotional facial expressions more negatively than controls (Bourke et al., 2010; Dalili et al., 2015) which may disrupt successful social interaction (Collin et al., 2013). Neurocognitive models suggest that emotion recognition biases may play a causal role in depression (Harmer et al., 2009). In addition to traditional medical treatment and therapy, addressing emotion recognition bias could help mitigate the long-term impact of depression and prevent further decline.

Several cognitive deficits have been found to be associated with depression, including impairments in working memory, visual memory and new learning (Burt & Zembar, 1995; Nikolin et al., 2021). Although the majority of studies have found that working memory is impaired in depression (Harvey et al., 2004; Rose & Ebmeier, 2006), some studies have shown that depression does not affect working memory capacity (Barch et al., 2003) or new learning (Taivalantti et al., 2020). Additionally, some cognitive deficits (e.g. attention, long term memory, working memory) can persist in remitted depression (Douglas & Porter, 2009; Semkovska et al., 2019) and may hinder functional recovery (Baune et al., 2010; Jaeger et al., 2006). 2010; Jaeger et al., 2006). Overall, these findings highlight the complexity of cognitive deficits in depression, suggesting that while some impairments may persist even after remission, their long-term effects on mental health remain uncertain, warranting further investigation.

While many previous studies have examined the relationship between cognitive abilities, facial emotion recognition bias, and depression, they tend to be smaller-scale experimental studies (Airaksinen et al., 2004; Anderson et al., 2011; Csukly et al., 2009; Gohier et al., 2009). Here, we used data from the Avon Longitudinal Study of Parents and Children (ALSPAC) cohort, which provides longitudinal data across early adulthood. The ALSPAC data base allowed us to investigate cross-sectional and longitudinal associations between depression and cognitive performance and whether depression in early adulthood has a lasting impact on cognitive performance. Establishing the longitudinal as well as the cross-sectional associations between cognitive abilities, facial emotion recognition bias, and depression could be instrumental in refining early detection methods and improving intervention strategies, ultimately leading to more effective treatments.

Most studies of emotion recognition biases in depression have used tasks in which participants identify emotions on faces displaying varying intensities of the same emotion (e.g. Anderson et al., 2011; Gollan et al., 2010). In this study, we used an emotion recognition bias measure involving morphed faces between happy and sad, creating ambiguous expressions (Penton-Voak et al., 2012). This approach differs from many emotion recognition tasks, which assess bias through false alarms and hit rates, and allows us to directly measure bias under conditions of uncertainty when presented with ambiguous faces. Also, findings on which specific emotion recognition is impaired in depression have been mixed. Some studies have found that depression is related to increased bias in sadness recognition (Milders et al., 2010; Suddell et al., 2021). However, meta-analyses have demonstrated an emotion recognition bias towards anger, disgust, fear, happiness, and surprise, but not sadness (Dalili et al., 2015), and that depression is associated with negative evaluations of neutral faces compared to controls (Bourke et al., 2010). These findings highlight the complexity of emotion recognition biases in depression, underscoring the need for further research to clarify these relationships.

We hypothesised that higher depressive symptoms would be associated with poorer performance on cognitive tasks and increased negative emotion recognition bias at age 31-32. Similarly, we predicted higher anxiety levels would be associated with poorer performance on cognitive tasks and greater emotion recognition bias. Additionally, lower well-being would be linked to poorer performance on cognitive tasks and increased emotion recognition bias.

Longitudinally, we hypothesised that meeting depression criteria at age 24 would predict poorer cognitive performance and increased negative bias in emotional judgements at ages 31-32. Additionally, we expected that, at age 24, low accuracy in recognising happy and sad faces, as well as overall accuracy in face recognition, would predict higher depressive symptoms at ages 31-32.

Additionally, we examined emotion recognition bias and cognitive performance in individuals with remitted depression to determine whether these effects persist after depressive symptoms subside. We hypothesised that individuals in remission would show a greater negative bias in emotional judgements and poorer cognitive performance compared to those who had not experienced high depressive symptoms, but less bias than those with current depression.

## Method

### Participants

We used The Avon Longitudinal Study of Parents and Children (ALSPAC) which is a cohort study that recruited children born in 1991 and 1992 in the former UK county of Avon (Boyd et al., 2013; Fraser et al., 2013; Northstone et al., 2019). The initial cohort consisted of 14,542 pregnant women, resulting in 14,062 live births, of which 13,988 children were alive at 1 year. An additional 913 eligible children were enrolled at the age of 7. Data collection occurred at regular intervals through questionnaires and in-clinic assessments. For this study, we included data from participants aged 31 and 32 years, collected during the 2021-2022 period, as well as data from the Focus@24 clinic when participants were 24 years old. Study data were collected and managed using REDCap electronic data capture tools hosted at the University of Bristol.1 REDCap (Research Electronic Data Capture) is a secure, web-based software platform designed to support data capture for research studies (Harris et al., 2009). This subset provided both cross-sectional and longitudinal perspectives on the participants’ health and cognitive abilities. We included all participants who provided data on at least one of the relevant measures across the study time points. Participants with missing data across all measurement points were excluded. For those with partial missing data, multiple imputation was used to handle missingness (see Missing data subsection for details). The final sample included 3,991 participants.

Ethical approval for the study was obtained from the ALSPAC Ethics and Law Committee and the Local Research Ethics Committees. Informed consent for the use of all data collected was obtained from participants following the recommendations of the ALSPAC Ethics and Law Committee at the time. Participants can contact the study team at any time to retrospectively withdraw consent for their data to be used. Study participation is voluntary and during all data collection sweeps, information was provided on the intended use of data. The completion of a questionnaire, either on paper or online, was considered to be written consent from participants to use their data for research purposes.

### Measures

A variety of cognitive and mental health data were collected from participants at these two data points (age 24 and age 31-32). The full data dictionary is available on the <u>ALSPAC</u> <u>study website</u>. For the timeline of the variables see Figure 1. At age 24, depression was assessed using the Revised Clinical Interview Schedule (CIS-R -Lewis et al., 1992), and emotion recognition was evaluated with the Emotion Recognition Task (ERT -Bamford et al., 2015). At ages 31–32, depression was measured using the Moods and Feelings Questionnaire (MFQ - Angold et al., 1995), well-being was assessed with the Warwick-Edinburgh Mental Well-being Scale (WEMWBS - Tennant et al., 2007), and anxiety was evaluated using the Generalised Anxiety Disorder Assessment (GAD-7 - Spitzer et al., 2006). Emotion recognition bias was assessed with the Emotion Bias Task (EBT - Penton-Voak et al., 2012), working memory capacity was measured with the Spatial Working Memory task (SWM - Huppert et al., 1995), and visual memory was evaluated using the Paired Associates Learning task (PAL Huppert et al., 1995). Further details on the measures used are available in the supplementary materials.

**Figure 1:**
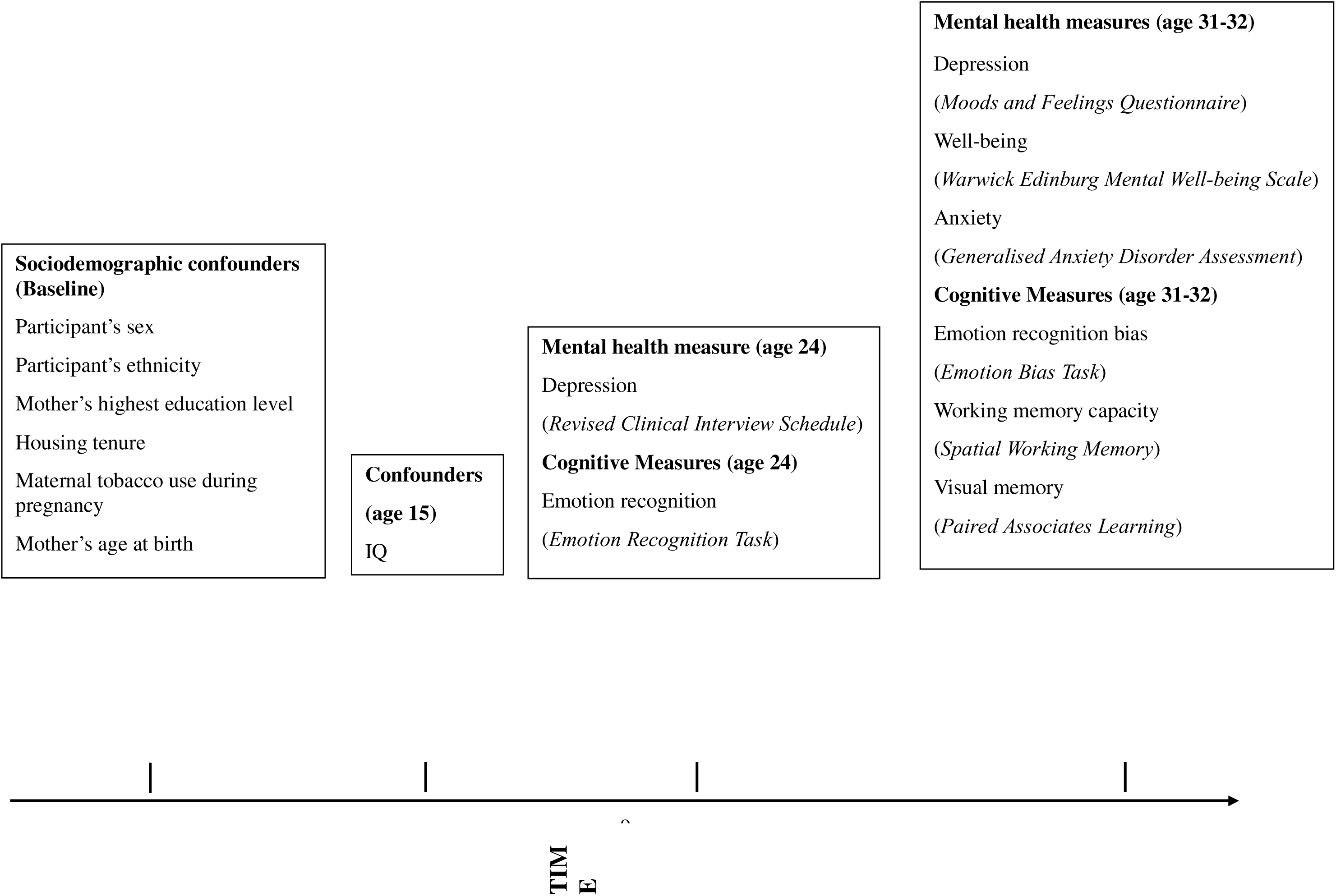
Timeline of ALSPAC variables used in the cross-sectional and longitudinal analyses.

#### Creating Remission Variable

To assess the effect of depression remission on emotion recognition bias and cognitive abilities, we created four new groups based on the CIS-R at age 24 and MFQ data at ages 31-32. We converted the MFQ into a binary variable using a cutoff point of 12 which indicates a high level of depressive symptoms, where 0 indicates no depression and 1 indicates the presence of depression. The CIS-R was already a binary variable. Group 1: Participants with low levels of depression at both time points. Group 2: Participants who did not meet the criteria for depression at age 24 but had high depressive symptoms at ages 31-32 (current depression). Group 3: Participants with high depressive symptoms at age 24 but not at ages 31-32 (remitted depression). Group 4: Participants who had high depressive symptoms at both time points. The remission variable was treated as a categorical predictor with Group 1 (no depression at both time points) as the reference category. Additionally, we created a binary depression variable to distinguish between participants who did not experience depression at these time points and those who had experienced depression at some point, including both time points.

#### Potential confounders

We adjusted for several sociodemographic variables known to be associated with mental health and cognitive abilities, including sex, maternal education level (determined during pregnancy and dummy coded as having O-levels or not), housing tenure (owned/mortgaged vs. other), maternal tobacco use during pregnancy (present/absent), mother’s age at birth, and child ethnicity (coded as non-white/white). To account for cognitive confounders, we included IQ, measured with the Wechsler Abbreviated Scale of Intelligence (Wechsler, 1981) at age 15, which was the nearest available time point to the exposures.

### Statistical analyses

Multiple imputation was conducted in RStudio version 4.1.2 with the Mice package (Buuren & Groothuis-Oudshoorn, 2011). We employed linear regressions to examine the relationships between depression, anxiety, and well-being, with emotion recognition bias and cognitive abilities. These relationships were assessed both cross-sectionally and longitudinally. We reported models both unadjusted and adjusted for confounders. The longitudinal models investigated whether depression at age 24 predicted emotion recognition bias and cognitive abilities at ages 31-32 and whether emotion recognition bias at age 24 predicted depression at age 31-32. The emotion recognition task (ERT) at age 24 differed from the emotional bias task (EBT) task at 31-32, as it assessed each emotion separately. We focused on the recognition of happy and sad emotions, as well as the total score, which consisted of recognition across six emotions: happy, sad, disgust, surprise, anger, and fear. Depression at ages 31-32 was measured using the MFQ. The cross-sectional models explored the associations between depression, anxiety, and well-being with emotion recognition bias and cognitive abilities.

For the emotion recognition bias and cognitive measures, data were collected at two time points: 2021 and 2022 (ages 31-32). There were 585 participants at the 2021 data point and 674 participants at the 2022 data point, with 174 participants providing data for both years. We combined these data points and used the first data point for each participant to mitigate potential training effects. For the mental health data collected at ages 31-32, there were five data collection points between 2020 and 2022. We selected the 2021 data point, as it had the most comprehensive dataset with the least amount of missing data.

The initial analyses were conducted without adjustments, while the second analyses were adjusted for confounders: participant’s IQ, sex, ethnicity, housing tenure, mother’s highest education level, mother’s age at birth, and mother’s tobacco use during pregnancy. We reported unstandardised regression coefficients (*b*) and standardised beta (β) values throughout the results using the imputed data sets. These longitudinal and cross-sectional analyses were conducted using the imputed datasets, with similar results observed in the non-imputed data. Full details are provided in the supplementary materials.

#### Missing data

We imputed the missing data because missing data can cause biased estimates and loss of statistical power in complete case analyses (Sterne et al., 2009). In this dataset, there was a high level of missingness (up to 70%) at the second data point (see supplementary materials for details Table 1). However, we chose to impute the missing data, as proper auxiliary variables can support accurate imputation even in cases of substantial missingness (Madley-Dowd et al., 2019). We created 100 imputed data sets using 10 iterations. We ran the analyses (described below) in each imputed data set and combined the results using Rubin’s rules (Rubin, 1987). We used multiple imputation for the main analyses, and results based on the non-imputed data are presented in the supplementary materials along with information on the missing data assessment, assumed relationship between variables and missing data indicators, auxiliary variables and imputation models. In selecting auxiliary variables, we prioritized those from the closest time points with the least missing data (Madley-Dowd et al., 2025). Continuous variables were imputed using predictive mean matching (PMM), while binary variables were imputed using logistic regression.

**Table 1.** Descriptive statistics for complete cases (N = 3991)

| **Variable** | **N (%) / Mean ± SD** |
| --- | --- |
| **Sex** (*Female*) | 2501 (62.7) |
| **Ethnicity** (*White*) | 2817 (70.6) |
| **Mothers social class** (*Unskilled*) | 66 (2.1) |
| *ii. Partly skilled* | 400 (12.5) |
| *iii. Skilled manual* | 111 (3.5) |
| *iv. Skilled non-manual* | 1312 (41.0) |
| *v. Managerial and technical* | 1138 (35.6) |
| *vi. Professional* | 174 (5.4) |
| **Depression diagnoses at 24** | 432 (10.9) |
| **Anxiety diagnoses at 24** | 385 (9.7) |
| **Mean depression score at 31-32 (SD)** | 4.50 (5.06) |
| **Mean anxiety score at 31-32 (SD)** | 5.54 (5.08) |
| **Mean well-being score at 31-32 (SD)** | 44.92 (9.24) |
| **Mean IQ at 15 (SD)** | 96.59 (12.99) |

## Results

### Cross-Sectional Relationships Between Emotion Recognition Bias and Mental Health

We ran linear regression analyses to investigate the cross-sectional relationship between emotion recognition bias and depression, well-being, and anxiety at the age of 31-32 (see Table 2). Results showed that higher depressive symptoms were associated with increased negative bias in emotional judgements (lower EBT scores) *b* = −.025, 95% CI [−.042, −.009], *p* = .003. Higher well-being was associated with a positive emotion recognition bias (higher EBT scores) *b* = .016, 95% CI [.007, .026], *p* = .001. Higher anxiety levels were associated with increased negative emotion recognition bias (lower EBT scores) *b* = −.019, 95% CI [−.036, −.003], *p* = .023.

**Table 2.** Regression results investigating the association between emotion recognition bias and mood measures cross-sectionally.

|  | Exposure | Unadjusted |  |  | Adjusted |  |  |
| --- | --- | --- | --- | --- | --- | --- | --- |
| | | *b*[95% CI] | $\beta$ | *p* | *b*[95% CI] | $\beta$ | *p* |
| **ER**<br>**Bias** | Depression | -.018[-.034, -.003] | -.070 | .023 | -.025[-.042, -.009] | -.097 | .003 |
|  | Well-being | .012[.003, .021] | .083 | .010 | .016[.007, .026] | .112 | .001 |
|  | Anxiety | -.014 [-.030, .002] | -.054 | .078 | -.019 [-.036, -.003] | -.075 | .023 |

*Notes: Emotion recognition bias towards morphed happy and sad facial expressions was measured with the Emotional Bias Task (EBT) at the age of 31-32. Higher scores on the EBT task indicate positive emotion recognition bias. Depression severity was measured with the MFQ at the age of 31-32. Higher scores on the MFQ suggest more severe depressive symptoms. Well-being was measured with the WEMWS at the age of 31-32. Higher scores of WEMWS indicate better well-being. Anxiety symptoms were measured with the GAD-7 at the age of 31-32. Higher scores in GAD-7 indicate higher anxiety symptoms. Adjustments included the participant's IQ, sex, ethnicity, housing tenure, mother's highest education level, mother's age at birth and mother's tobacco use during pregnancy.*

### Cross-Sectional Relationships Between Cognitive Performance and Mental Health

We ran linear regression analyses to investigate the cross-sectional relationship between cognitive performance and depression, well-being, and anxiety. No association was found between working memory or visual memory and depression, well-being or anxiety (see Table 3).

**Table 3.**
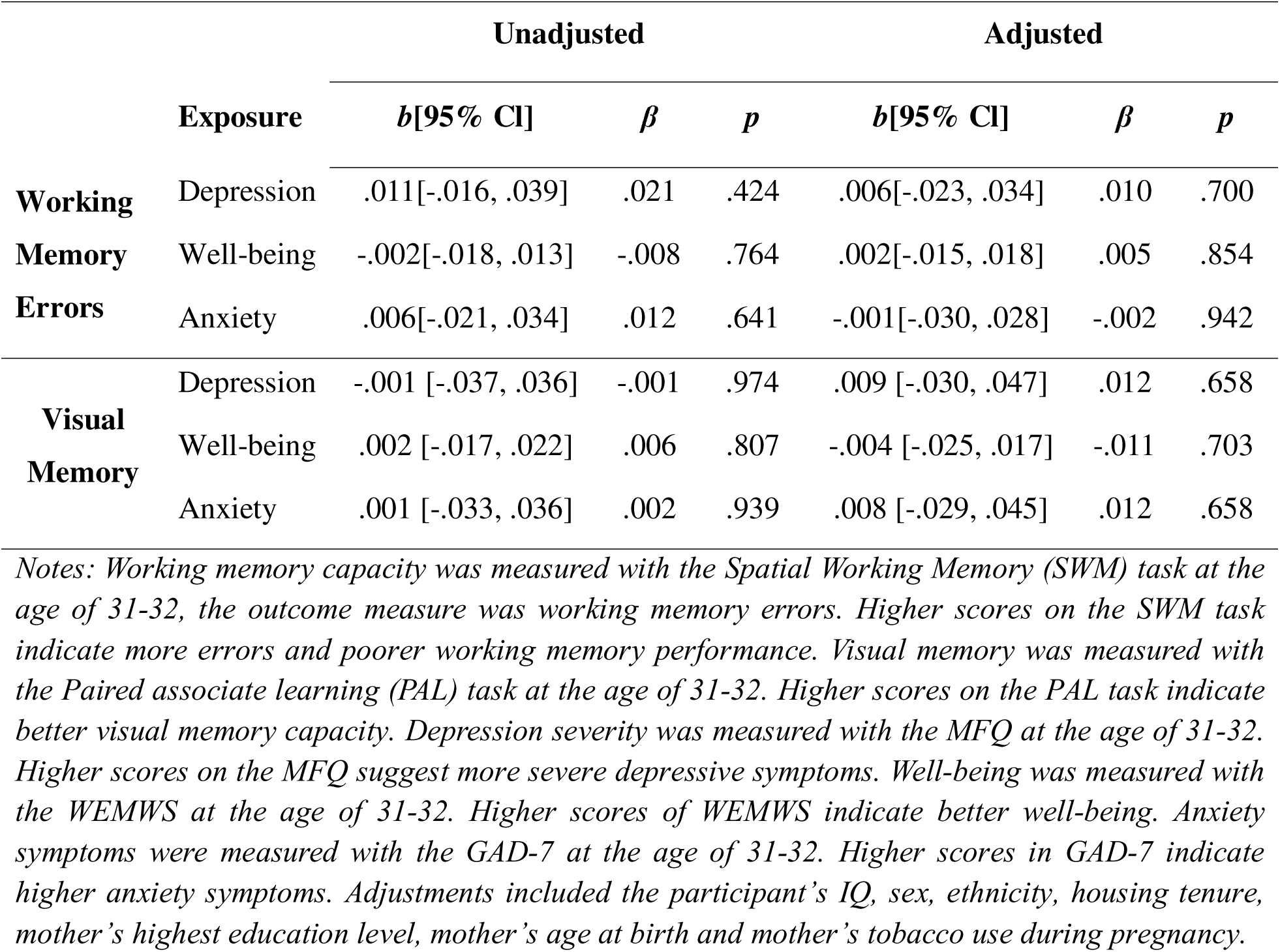
Regression results investigating the association between cognitive measures and mood measures cross-sectionally.

### Longitudinal Relationships Between Emotion Recognition Bias, Cognitive Performance and Depression

We ran linear regression analyses to investigate the longitudinal relationships between emotion recognition bias, cognitive performance, and depression (see Table 3). Results showed that depression at the age of 24 predicted higher negative emotion recognition bias at the age of 31-32, in the adjusted model, *b* = −.362, 95% CI [−.630, −.094], *p* = .008. There was no evidence that depression predicted working memory or visual memory performance at the age of 31-32 (see Table 4).

**Table 4.**
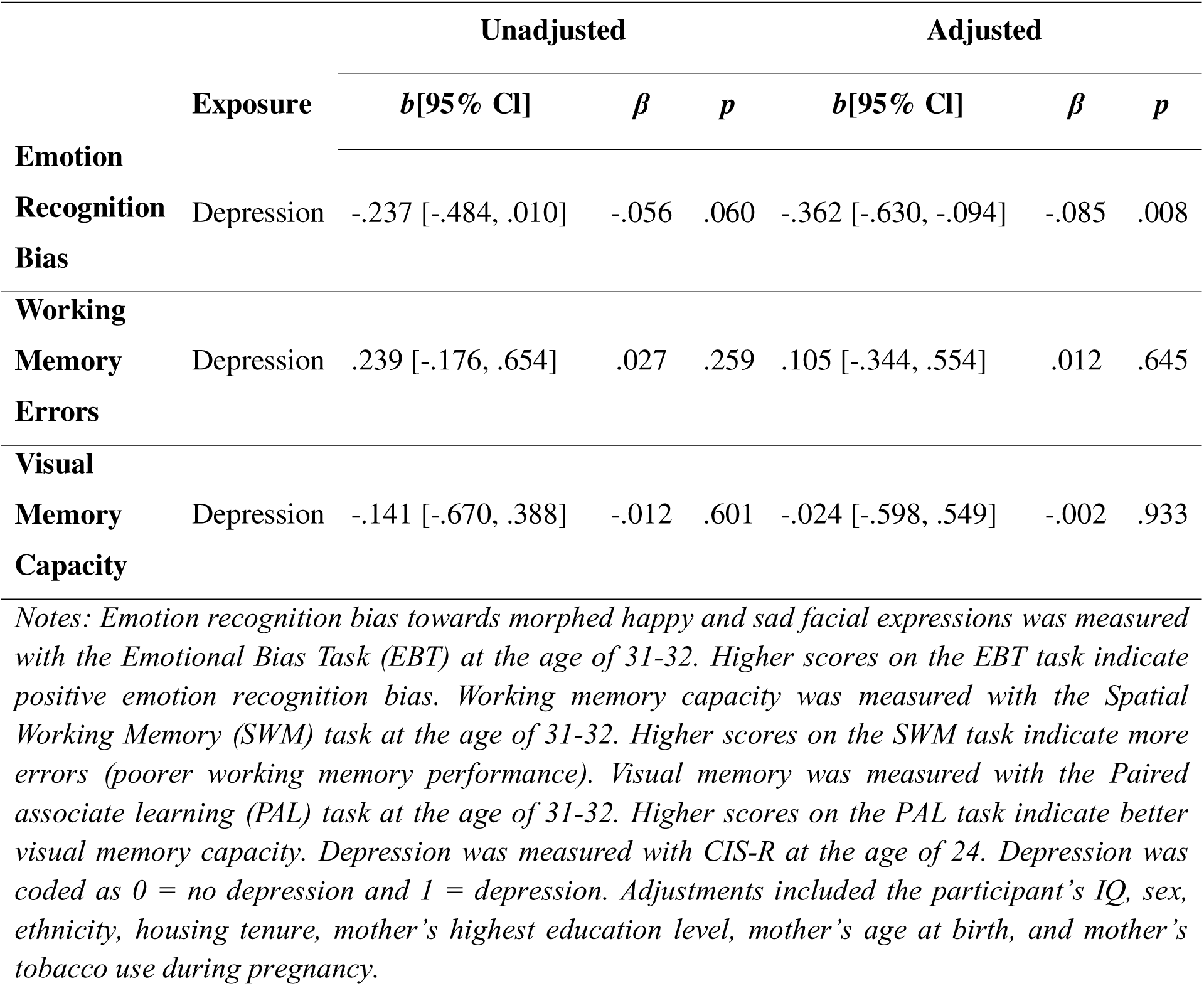
Regression results investigating the association between emotion recognition bias, cognitive measures and depression longitudinally.

We also explored the longitudinal relationships between emotion recognition ability and depression, this time in the reverse direction. We examined whether performance on the emotion recognition task at age 24 predicted depression at ages 31-32. Results showed that reduced accuracy in recognising happy emotions predicted higher depressive symptoms *b* = −.176, 95% CI [−.274, −.079], *p* <.001. There was no evidence that the accuracy of recognising sad faces or the total accuracy predicted depression at the age of 31-32 (see Table 5).

**Table 5.**
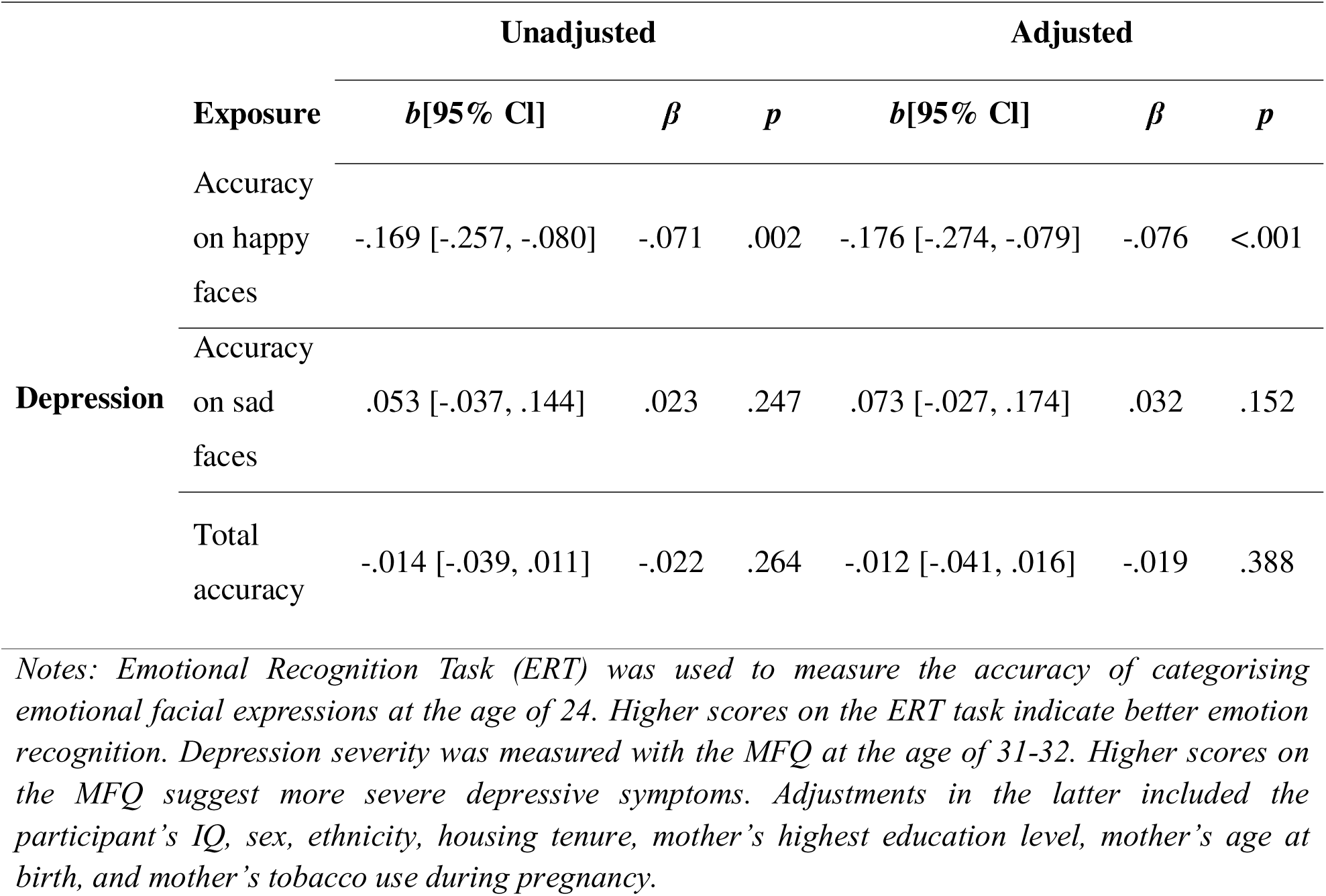
Regression results investigating the association between accuracy of emotion recognition and depression longitudinally.

**Table 6.** Descriptive statistics of emotion recognition bias scores for each depression status group.

| **Depression Status** | **Mean** | **SD** | **N** |
| --- | --- | --- | --- |
| 1 (No depression at both time points) | 7.32 | 1.34 | 2481 |
| 2 (Current depression) | 7.19 | 1.39 | 965 |
| 3 (Remitted depression) | 7.12 | 1.37 | 297 |
| 4 (Depression at both time points) | 7.10 | 1.43 | 247 |

### Relationships Between Emotion Recognition Bias, Cognitive Performance and Depression Remission

A regression analysis was conducted to examine the relationship between remission status and emotion recognition bias. There was no evidence that current high depressive symptoms, remitted depression, or high depressive symptoms at both time points affected emotion recognition bias when compared to lower depressive symptoms (see Table 7). Results indicated weak evidence that having high depressive symptoms at any point was associated with a higher negative emotion recognition bias compared to those who did not have depression at both time points. However, this effect weakened after adjusting for confounders, *b* = −.10, 95%CI [−.24, .04], *p* = .16.

**Table 7.**
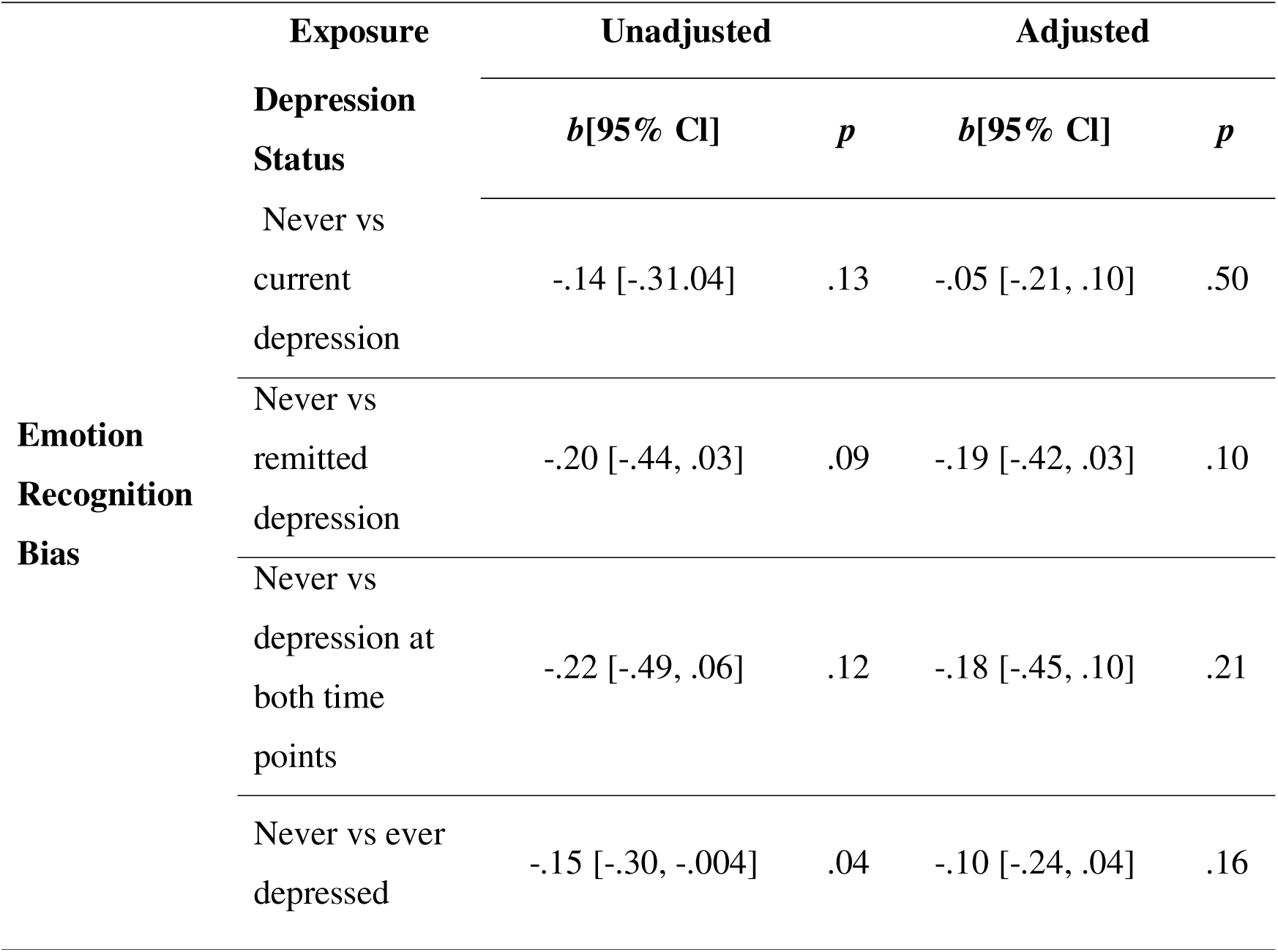
Regression results of emotion recognition bias scores for each depression status group.

**Table 8.**
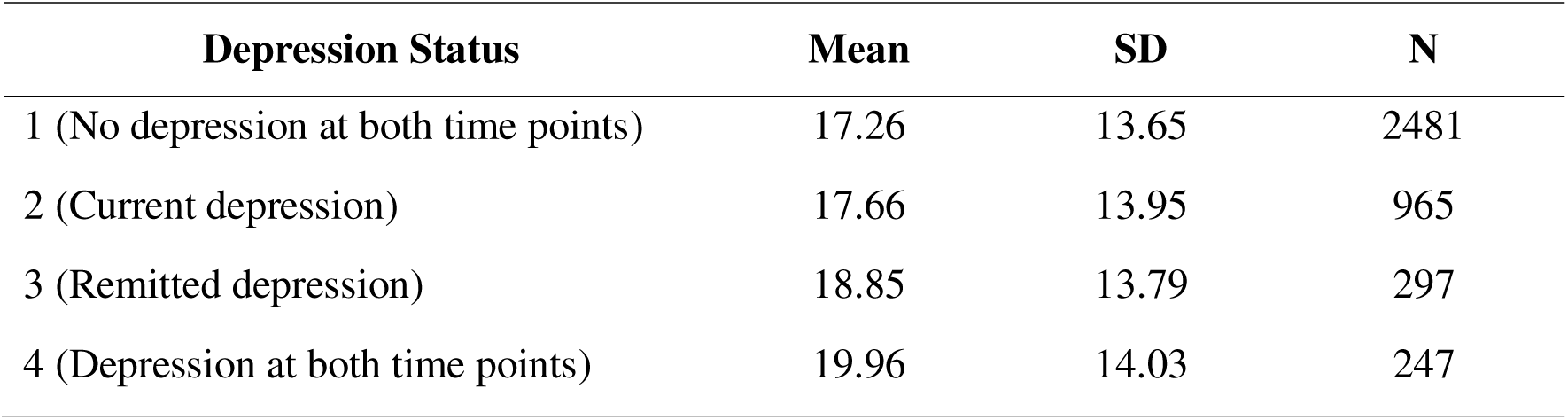
Descriptive statistics of working memory error scores for each depression status group.

| **Depression Status** | **Mean** | **SD** | **N** |
| --- | --- | --- | --- |
| 1 (No depression at both time points) | 17.26 | 13.65 | 2481 |
| 2 (Current depression) | 17.66 | 13.95 | 965 |
| 3 (Remitted depression) | 18.85 | 13.79 | 297 |
| 4 (Depression at both time points) | 19.96 | 14.03 | 247 |

A regression analysis was conducted to assess the relationship between remission status and working memory errors. There was no evidence that current high depressive symptoms, remitted depression, or high depressive symptoms at both time points affected working memory performance when compared to lower depressive symptoms (see Table 9).

**Table 9.**
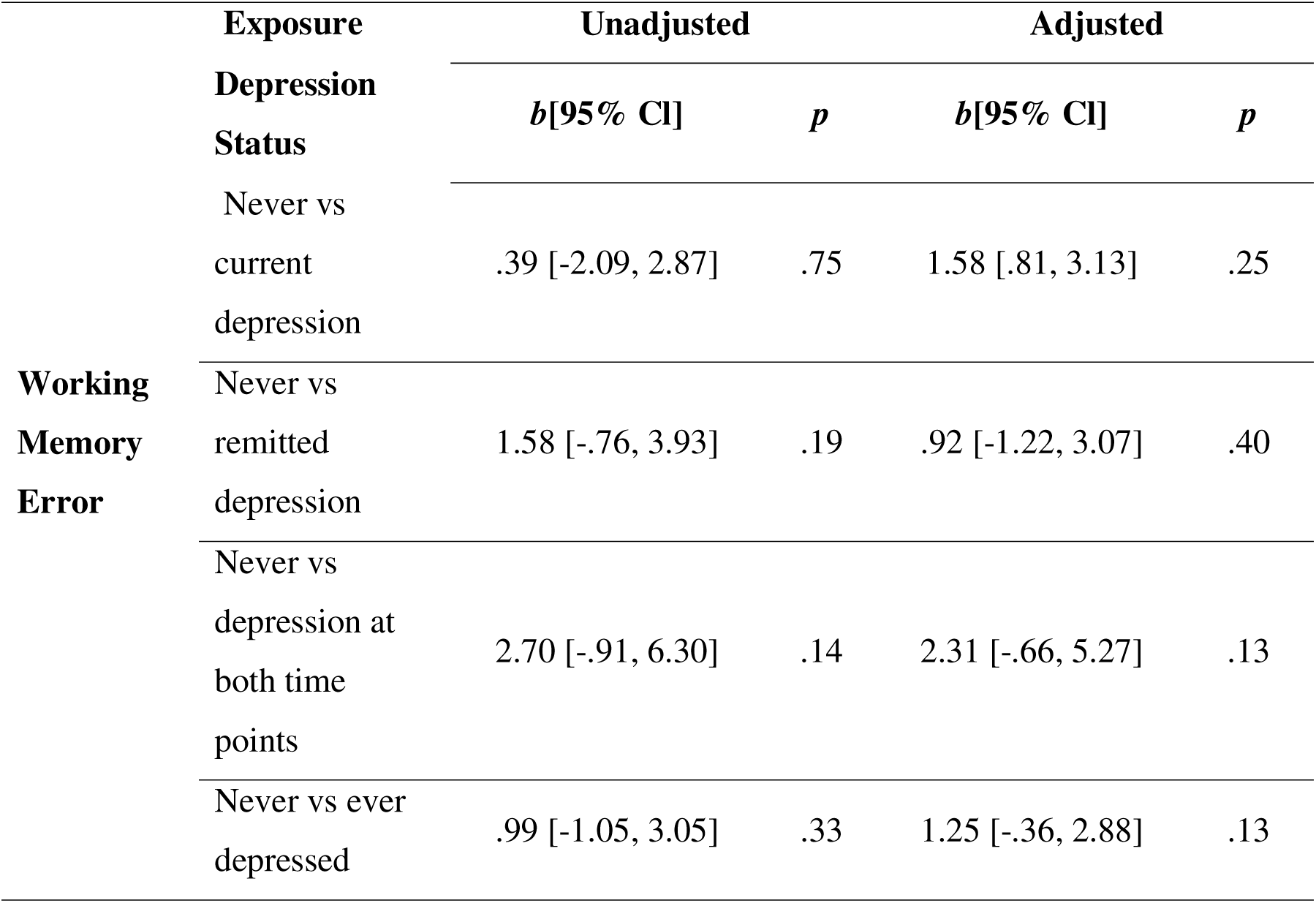
Regression results of working memory error scores for each depression status group.

## Discussion

We investigated both cross-sectional and longitudinal associations between depression, cognitive performance, and emotion recognition bias, and we also explored the potential impact of anxiety and well-being on these outcomes. Results indicated that there was some evidence higher depressive symptoms were associated with greater negative emotion recognition bias in cross-sectional analyses. Additionally, meeting the criteria for depression predicted later negative emotion recognition bias, aligning with existing literature (Bourke et al., 2010; Krause et al., 2021). Notably, reduced accuracy of recognising happy faces, but not sad faces, predicted future depressive symptoms, aligning with research suggesting that recognising happy faces is impaired in individuals with depression (Dalili et al., 2015). Some evidence also suggested that lower well-being and higher anxiety symptoms were related to negative emotion recognition bias cross-sectionally, while the existing literature on this relationship has yielded mixed findings (Dyer et al., 2021; Surcinelli et al., 2006). Our results suggest that negative emotion recognition bias is related to overall poorer mental health rather than being specific to depression.

Cross-sectional analyses and longitudinal analysis showed no evidence depression was associated with working memory performance, contradicting some previous findings (Harvey et al., 2004). There was no evidence of a relationship between visual memory and depression, either cross-sectionally or longitudinally, or with well-being or anxiety, aligning with previous findings showing no association between depression and performance on the visual PAL task (Taivalantti et al., 2020). This may be because the PAL task was primarily developed to detect memory deficits in dementia (Barnett et al., 2015). It is possible that the task was too easy for this participant group, despite its varying difficulty levels; however, the distribution of the data did not indicate a clear ceiling effect.

Results suggested that emotion recognition bias persists over time, and this finding was consistent with a causal role for emotion perception biases in depression (Harmer et al., 2009). To further investigate this, we examined depression remission to assess how remitted and current depression relate to working memory performance and emotion recognition bias. The results showed no evidence that the state of depression, whether current or remitted, differed from non-depression in terms of emotion recognition bias or working memory performance. These findings do not support previous research which found that participants in remission from depression were better at recognizing negative facial expressions compared to healthy controls (Anderson et al., 2011). However, there was weak evidence suggesting that experiencing depression, whether in the past or currently, was associated with greater negative emotion recognition bias. However, caution is needed when interpreting these results, as the sample sizes for the different groups were unevenly distributed, with low numbers in the depressed and remitted groups.

This study has potential clinical implications, as the results showed some cross-sectional and bi-directional longitudinal relationships between emotion recognition bias and depression. These findings support previous research suggesting that emotion recognition biases may contribute to the onset and maintenance of depression (Harmer et al., 2009). Combined with existing evidence, these results further support the idea that targeting emotion recognition bias could be a potential intervention strategy for reducing depressive symptoms. However, we did not find strong longitudinal associations between other aspects of cognitive performance and depression, despite observing some cross-sectional associations in the unadjusted models. This suggests that emotion recognition bias may be more persistent than cognitive performance decline. Targeting emotion recognition bias may be more beneficial for long-term intervention and prevention of recurrence. It is important to note that this interpretation assumes that emotion recognition biases lie on the causal pathway of depression—a hypothesis that was not directly tested in this study, though our results do strengthen the evidence supporting this possibility (Harmer et al., 2009).

One of the strengths of this study was its use of a large longitudinal dataset, which provided greater statistical power compared to smaller-scale experimental studies. This enhances the ability to detect small effects, which are often prevalent in mental health research (Dalili et al., 2015). The longitudinal nature of the data provided valuable insights into the evolving relationships between depression, cognitive abilities, and emotion recognition bias over time. Such longitudinal analysis is critical for identifying patterns and causal pathways that might be missed in cross-sectional studies, offering a more comprehensive view of the long-term impact of depression on cognitive and emotional functioning.

One limitation of this study was the difference in depression measures used across time points. At age 24, depression was assessed using a clinical measure with a binary outcome (presence or absence of depression), whereas at ages 31-32, continuous measures of symptom severity were employed. A similar limitation applies to the emotion recognition measures. At age 24, participants completed a task assessing their ability to recognise different emotions presented at varying intensities. In contrast, at ages 31-32, the task specifically measured emotion recognition bias, using morphed faces between happy and sad to assess bias towards ambiguous emotional expressions. Although these tasks may appear similar, they differ in focus: the emotion recognition task measured participants’ ability to identify individual emotions, whereas the emotion recognition bias task specifically examined bias towards happy or sad expressions These differences in measurement hinder direct comparison between cross-sectional and longitudinal results. Additionally, since the study participants were from the general population rather than being clinically diagnosed with depression, the findings may not fully capture the experiences of individuals with more severe or chronic depressive disorders. Another limitation is that some measures used to assess depression may not be as accurate as others. For example, the measure used at age 24, the CIS-R, is a clinical tool. However, the MFQ cut-off point of 12, which we used in the remission data, may not be as precise as the CIS-R. We chose to use multiple imputation because complete case analysis is likely to be biased, as the missing data appear to depend on the outcome (Hughes et al., 2019). Additionally, there are limitations associated with the imputation method employed. We used multiple imputation, which assume that data are missing at random. However, in this study, missingness may be missing not at random, as individuals with higher levels of depression at the time of assessment might have been less likely to participate. While multiple imputation may also be biased if the data are not missing at random, we included auxiliary variables in the imputation model to make the missingness at random assumption more plausible and reduce potential bias.

Future studies should explore the effects of treated versus untreated depression to determine whether the impacts on cognitive abilities and emotion recognition bias are reversible. Given that emotion recognition bias was both predicted by and predictive of depression over time, future research should investigate the effectiveness of interventions targeting emotion recognition bias in reducing depression severity or preventing recurrence.

In conclusion, this study demonstrated that depression in young adulthood predicts later emotion recognition bias, and emotion recognition bias in turn predicts future depressive symptoms. The study also revealed cross-sectional associations between emotion recognition and depression, as well as with anxiety and well-being. We did not observe any longitudinal associations of depression on working memory and no relationship was found between depression and visual memory. We found no evidence that remitted and non-remitted depression differed in terms of emotion recognition bias and cognitive performance, though our sample lacked sufficient variability to assess this thoroughly. These findings highlight emotion recognition bias as an important target for intervention, in line with existing research. Future research should investigate whether modifying emotion recognition bias can reduce the risk of future depressive episodes.

## Acknowledgements

We are extremely grateful to all the families who took part in this study, the midwives for their help in recruiting them, and the whole ALSPAC team, which includes data collection staff, data and administrations staff, technical managers and the technical staff with the Bristol Bioresource Laboratory, based within the University of Bristol.

## Data availability statement

The informed consent obtained from ALSPAC (Avon Longitudinal Study of Parents and Children) participants does not allow the data to be made available through any third party maintained public repository. Supporting data are available from ALSPAC on request under the approved proposal number, B4245. Full instructions for applying for data access can be found here: http://www.bristol.ac.uk/alspac/researchers/access/. The ALSPAC study website contains details of all available data (http://www.bristol.ac.uk/alspac/researchers/our-data/).

## AI Use Statement

Artificial intelligence tools were used to assist in data management, script development, code formatting, and performing grammar and spelling checks. The authors reviewed and verified all outputs to ensure accuracy and integrity.

## Supplementary Materials

### Measures

#### Mood Measures

##### Revised Clinical Interview Schedule

At the age of 24 years, participants completed the Revised Clinical Interview Schedule (CIS-R) which is a self-report questionnaire that assesses a range of neurotic symptoms related to depression and anxiety (Lewis et al., 1992). We utilized this depression variable, which captured primary or secondary diagnoses of mild, moderate, or severe depression. To fit our regression model, we dummy-coded this variable into a binary format, distinguishing between meeting the criteria for depression and not meeting the criteria for depression.

##### Moods and Feelings Questionnaire (MFQ)

At ages 31-32 years, depression symptoms were measured using the Moods and Feelings Questionnaire (MFQ), which consists of 13 questions assessing how participants have felt or acted recently (Angold et al., 1995). This questionnaire uses a 3-point scale: “not true,” “sometimes true,” and “true.” Scores range from 0 to 2 for each item, with total scores ranging from 0 to 26. Higher scores on the MFQ indicate more severe depressive symptoms.

##### Warwick Edinburg Mental Well-being Scale (WEMWBS)

At ages 31-32 years, mental well-being was measured using the Warwick-Edinburgh Mental Well-being Scale (WEMWBS). This scale consists of 14 questions with each item scored from 1 (“none of the time”) to 5 (“all of the time.”), with total scores ranging from 14 to 70 (Tennant et al., 2007). Higher scores indicate higher well-being.

##### Generalised Anxiety Disorder Assessment (GAD-7)

At ages 31-32 years, anxiety was measured using the Generalized Anxiety Disorder 7-item Scale (GAD-7), which consists of 7 questions assessing generalized anxiety (Spitzer et al., 2006). This scale uses a 4-point response format: “not at all,” “several days,” “more than half the days,” and “nearly every day.” Scores range from 0 to 3 per item, with total scores ranging from 0 to 21. Scores of 5, 10, and 15 are used as cut-off points for mild, moderate, and severe anxiety, respectively. Higher scores on the GAD-7 indicate more severe anxiety symptoms.

#### Cognitive Measures

##### Emotion Recognition Task (ERT)

At the age of 24 years, participants completed the Emotion Recognition Task (ERT), which assessed their accuracy in recognizing facial emotional expressions(Bamford et al., 2015). This task was from the CANTAB battery (<u>Cambridge Cognition</u>). Participants were presented with six emotional expressions at different intensities: happiness, sadness, anger, disgust, fear, and surprise. The stimuli had eight levels of intensity, ranging from near-neutral expressions to prototypical displays. The primary outcome was the total number of correct identifications (“hits”). For this study, we focused only on the accuracy of happy, sad, and total recognition.

##### Emotion Bias Task (EBT)

At ages 31-32 years, emotion recognition bias was measured using the Emotion Bias Task (EBT). In this task, participants were shown images of faces and asked to determine whether each face was happy or sad. The facial images (15 in total) were morphed between happy and sad emotions, ranging from a fully happy face to a fully sad face, with 15 levels of intensity between these extremes. Each stimulus varied in the ratio of the two emotions presented. The primary outcome measure was the “balance point,” which represents the image at which a participant was equally likely to categorize the face as happy or sad. The balance point was calculated as the number of ‘happy’ responses divided by the total number of trials in the baseline/test block, and this number was then multiplied by the number of stimuli (Penton-Voak et al., 2012). A higher balance point indicates a lower bias toward emotional facial expressions.

##### Paired Associates Learning (PAL)

At ages 31-32 years, the Paired Associates Learning (PAL) task was used to assess visual memory and new learning (Huppert et al., 1995). This task was from the CANTAB battery (<u>Cambridge Cognition</u>). In this task, participants viewed a series of boxes that opened in random order, some of which contained patterns. Participants were required to select the boxes that contained the patterns shown in the middle of the screen. The primary outcome measures were PAL-total errors adjusted, which included the number of errors plus an adjustment for stages not reached, and PAL-errors in the six-shapes stage, which counted the errors made during the stage with six boxes. Higher scores indicate poorer performance in visual memory and learning.

##### Spatial Working Memory (SWM)

At ages 31-32, the Spatial Working Memory (SWM) task was used to assess visual working memory capacity. This task was from the CANTAB battery (<u>Cambridge Cognition</u>). This task involves participants viewing a series of coloured boxes on the screen and searching for a yellow token. The number of boxes on the screen gradually increases up to 12 (Huppert et al., 1995). The outcome measures include the number of errors (the number of times a participant revisited a box where a token had previously been found) and the strategy used (the number of times a participant started a new search by selecting a different box). Lower scores indicate better cognitive performance, as they reflect fewer errors and a more effective search strategy. For this analysis, we focused on the number of error boxes, with lower scores indicating better performance.

## Supplementary Materials Tables

**Table 1.** Missingness percentages of imputed and auxiliary variables.

| Imputed variables |  | Auxiliary variables |  |
| --- | --- | --- | --- |
| Variable | Missing Percentage | Variable | Missing Percentage |
| Remission | 31.55% | Nbackat_24 | 10.90% |
| remission_binary | 31.55% | Danva_happy_errors_at8 | 25.36% |
| MFQ_at32_binary | 31.20% | Danva_sad_errors_at8 | 25.36% |
| MFQ_at32 | 31.20% | Danva_angry_errors_at8 | 25.36% |
| GAD7_at32 | 30.44% | Danva_fear_errors_at8 | 25.36% |
| WEMWS_at32 | 29.07% | Danva_total_errors_at8 | 25.36% |
| Depression_at24 | 0.68% | IQ_at8 | 19.77% |
| SWM_at32 | 77.88% | WM_at10 | 22.28% |
| EBT_at32 | 77.88% | SST_at24 | 14.06% |
| PAL_at32 | 77.77% | Anxiety_at24 | 0.93% |
| Happycorrect_at24 | 9.52% | Angrycorrect_at24 | 9.52% |
| Sadcorrect_at24 | 9.52% | Disgustcorrect_at24 | 9.52% |
| Totalcorrect_at24 | 9.52% | Fearcorrect_at24 | 9.52% |
| sex | 0.00% | Surprisecorrect_at24 | 9.52% |
| IQ_at15 | 25.96% | car_ownership | 20.27% |
| ethnicity | 0.00% |  |  |
| mothers_smoking | 12.60% |  |  |
| mothers_age_atbirth | 15.99% |  |  |
| housing_tenure | 43.67% |  |  |
| mothers_social_class | 19.79% |  |  |

**Table 2.** Cross-Sectional Relationship Between Emotion Recognition Bias and Mental Health.

|  | Exposure | Unadjusted |  | Adjusted |  |
| --- | --- | --- | --- | --- | --- |
|  |  | *b*[95% CI] | *p* | *b*[95% CI] | *p* |
| **Emotion**<br>**Recognition**<br>**Bias** | Depression | -.02[-.03, -.0003] | .045 | -.03[-.06, -.01] | .008 |
|  | Well-being | .01[.001, .02] | .03 | .02[.01, .04] | <.001 |
|  | Anxiety | -.01[-.03, .001] | .06 | -.03[-.05, -.005] | .02 |

*Notes: Emotion recognition bias towards morphed happy and sad facial expressions was measured with the Emotional Bias Task (EBT) at the age of 31-32. Higher scores on the EBT task indicate positive emotion recognition bias. Depression severity was measured with the MFQ at the age of 31-32. Higher scores on the MFQ suggest more severe depressive symptoms. Well-being was measured with the WEMWS at the age of 31-32. Higher scores of WEMWS indicate better well-being. Anxiety symptoms were measured with the GAD-7 at the age of 31-32. Higher scores in GAD-7 indicate higher anxiety symptoms. N(MFQ) = 1055. N(WEMWS) = 1014. N(GAD7) = 1011. N(WEMWS) = 1014. Adjustments in the latter included the participant's IQ, sex, ethnicity, housing tenure, mother's highest education level, mother's age at birth and mother's tobacco use during pregnancy.*

**Table 3.**
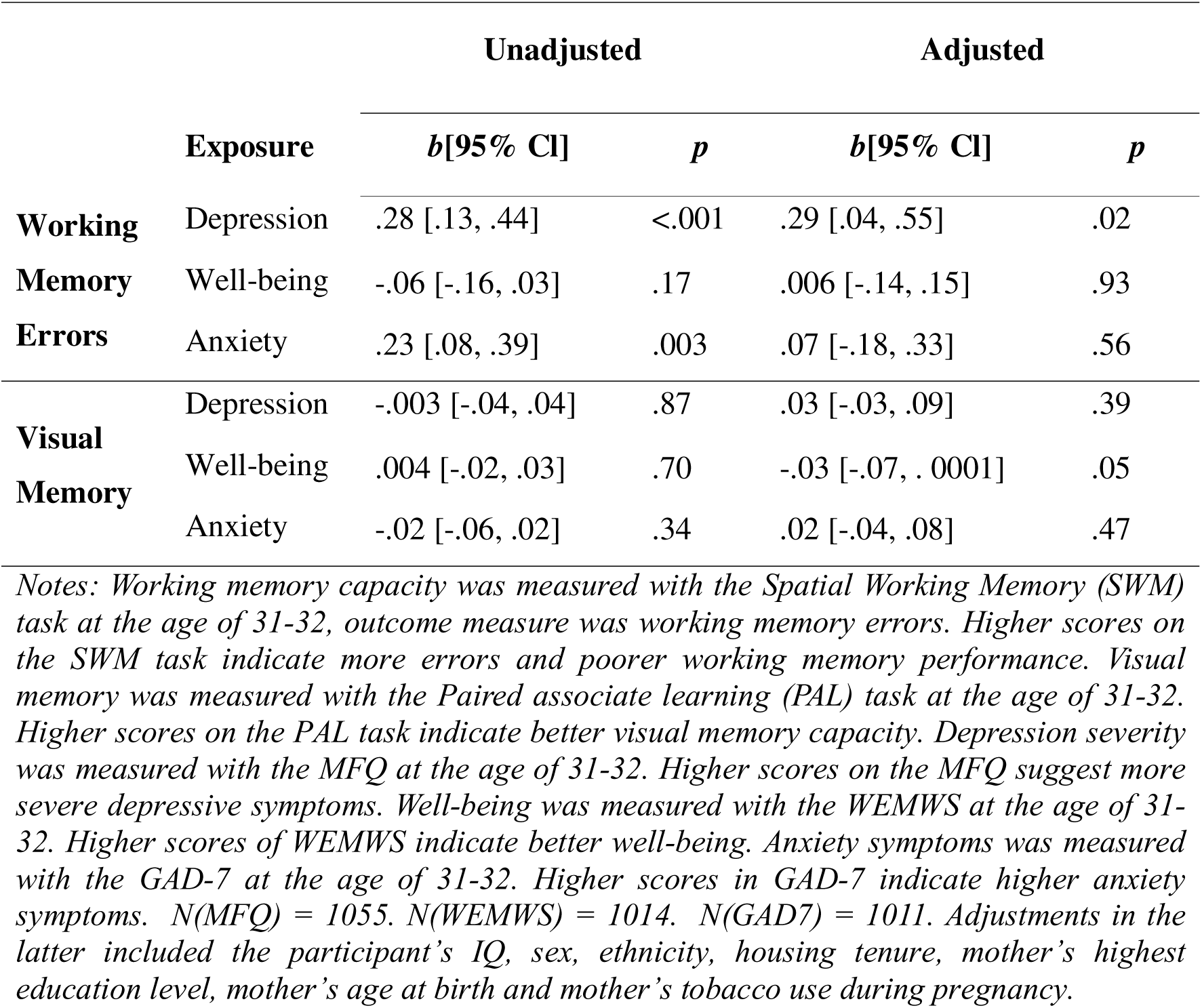
Cross-Sectional Relationship Between Cognitive Performance and Mental Health (Non-imputed data)

**Table 4.** Longitudinal Relationships Between Emotion Recognition Bias, Cognitive Performance and Depression (Non-imputed data)

|  | Exposure | Unadjusted |  | Adjusted |  |
| --- | --- | --- | --- | --- | --- |
|  |  | *b*[95% CI] | *p* | *b*[95% CI] | *p* |
| **Emotion Recognition Bias** | Depression | -.29 [-.58, -.0001] | .049 | -.82 [-.38, -1.25] | <.001 |
| **Working Memory Errors** | Depression | 2.63 [-.34, 5.60] | .08 | 1.24 [-3.21, 5.71] | .58 |
| **Visual Memory Capacity** | Depression | -.30 [-1.04, .45] | .43 | -.54 [-1.61, .54] | .33 |

*Notes: Emotion recognition bias towards morphed happy and sad facial expressions was measured with the Emotional Bias Task (EBT) at the age of 31-32. Higher scores on the EBT task indicate positive emotion recognition bias. Working memory capacity was measured with the Spatial Working Memory (SWM) task at the age of 31-32. Higher scores on the SWM task indicate more errors and poorer working memory performance. Visual memory was measured with the Paired associate learning (PAL) task at the age of 31-32. Higher scores on the PAL task indicate better visual memory capacity. Depression was measured with CIS-R at the age of 24. Depression was coded as 0 = no depression and 1 = depression. N(EBT) = 879. N (SWM) = 878. N(PAL) = 883. Adjustments in the latter included the participant's IQ, sex, ethnicity, housing tenure, mother's highest education level, mother's age at birth, and mother's tobacco use during pregnancy.*

